# PrEP Engagement and Youth Experiencing Homelessness in a sample of Youth Experiencing Homelessness in New York City: A Brief Report

**DOI:** 10.1101/2024.10.23.24315874

**Authors:** Amanda Sisselman-Borgia, Jonathan Ross, Dana Watnick, Nicole Saint-Louis

**Affiliations:** Lehman College Department of Social Work, 250 Bedford Park Blvd West, Bronx NY 10468; Division of General Internal Medicine, Albert Einstein College of Medicine/Montefiore Medical Center, 4234 Bronx Blvd, Bronx, NY 10466; Albert Einstein College of Medicine, Department of Pediatrics, 1225 Morris Park Avenue, Van Etten 6B23, Bronx, NY 10461

## Abstract

Few studies have examined pre-exposure prophylaxis (PrEP) engagement among youth experiencing homelessness (YEH), despite their high HIV risk. We assessed awareness of, willingness to use, and plans to take PrEP among YEH receiving services at nine organizations in New York City. Among 113 participants, 49 (43%) identified as LGBTQIA and 74 (65%) reported food insecurity. In total, 53 (47%) had heard of PrEP before the survey, 82 (73%) reported willingness to take PrEP, and 28 (25%) had a plan to take it. Interventions to improve awareness of PrEP and encourage use are critical to reducing HIV in YEH.

## Introduction

Nationally, over 4 million children and young adults annually are impacted by physical, mental, and emotional problems related to being unhoused,^1^ and this problem is growing: between 2015 and 2018 there was a 16% increase in youth experiencing homelessness (YEH). Homelessness is an independent predictor of both HIV infection and HIV-related mortality.^1^ Nearly 25% of new HIV infections occur in youth ages 13 to 24 years old and YEH are disproportionately impacted. While national data are lacking, up to 20% of YEH in New York City (NYC) - which has one of the largest populations of YEH in the U.S. - are estimated to be living with HIV.^2^ Thus, treatment and prevention of HIV among YEH is critical to ending the epidemic, particularly as rates of youth homelessness continue to rise.

Pre-exposure prophylaxis (PrEP) is a highly effective tool for HIV prevention, yet the limited available data suggest it is under-utilized among YEH.^3,4^ A study of over 1400 YEH in 7 US cities found that although 84% of those sampled were eligible for PrEP, only 29% knew what it was; nonetheless, 59% reported interest or willingness to use it.^4^ Multiple intersecting barriers exist impacting awareness of and access to PrEP for YEH, including anticipated stigmas related to HIV, homelessness, sexuality and gender; discrimination; lack of peer acceptance; and hesitancy among YEH to engage with medical providers.^4,5^ Medical providers are also sometimes hesitant to prescribe PrEP because of concerns about disinhibition and risk compensation.^6^

The majority of YEH in the US are Black, Indigenous, and other people of color (BIPOC), with more than 50% identifying as LGBTQ+; this proportion is even higher in NYC.^1^ Youth who identify as LGBTQ+ are more likely than straight youth to trade sex for food, housing, or other necessities and are also more likely to be diagnosed with HIV.^2,7^ Transgender youth are particularly at risk: nearly 75% experience periods of homelessness, and they are more likely than cis-gendered youth to engage in transactional sex and use injectable drugs, both of which increase HIV risk.^7^ Further, these same factors create challenges for health system delivery of HIV prevention interventions to this population, particularly PrEP.

To date, almost no studies have examined PrEP engagement among YEH, despite a clear need to understand and address their complex and intersecting determinants of health. To address this gap, we describe awareness of PrEP, willingness to use PrEP, and plans to take PrEP among a sample of YEH in NYC.

## Methods

### Setting and Participants

We conducted this study in NYC, where 69% of new infections in 2022 were in people under the age of 40.^2^ Homeless persons are disproportionately impacted, accounting for 10% of new HIV infections in 2022.^2^ There are approximately 8100 YEH in NYC on any given day, but the actual number of YEH is likely even higher than 8100, as this only reflects those that the city is able to find and count.^8^ Data for the current study were collected across 9 sites providing services for YEH in NYC in all five boroughs, including four transitional residences and one crisis shelter run by community providers through the NYC Department of Youth and Community Development (DYCD) shelter programs, as well as five drop-in centers designated by the NYC Department of Children and Youth as safe spaces to hang out, get food, shower and change clothes.

### Data Collection

Study recruitment fliers were distributed to sites identifying specific dates the study team would be on-site. Trained research assistants (RAs) met with groups of YEH at study sites, to describe the study and recruit survey participants. Participants were eligible to complete the survey if they were between the ages of 16 and 24 and spoke and read English fluently. Interested persons who were eligible received a document describing the study; RAs also explained the study to them in detail, including procedures and associated benefits and risks. Upon survey completion, participants were given a $20 gift card to a local fast food restaurant. The CUNY Institutional Review Board granted a waiver of signed consent for this study, such that completion of the anonymous survey was considered implied consent to participate.

### Measures and analysis

We examined three primary outcomes for this study. *Awareness of PrEP* (assessed by asking, “Have you heard of PrEP before taking this survey?), *willingness to use* PrEP (initially assessed on a 5-point Likert scale, but for analysis purposes, we dichotomized the interest variable into two options – interested/not interested), and *future plans to use PrEP* (assessed by asking, “PrEP is currently available with a prescription and is covered by most insurances in NY. Do you plan to take PrEP?”). ^4,9,10^ Sociodemographic variables collected included age, gender, race/ethnicity, sexual orientation, employment status, level of education, injection drug use (yes/no), prior transactional sex (yes/no), condomless sex (yes/no), and health insurance status (yes/no). Finally, we collected data about potential barriers and facilitators to PrEP engagement, including concerns about side effects and medication cost, self-assessed access to pharmacy or doctor, and concern about having to remember to take a pill each day.^4,9^ Participants were asked to ‘select all’ factors that might prevent them from obtaining and taking PrEP.

We used descriptive statistics to measure frequencies of demographic items, main outcome variables, and barriers to PrEP use. Chi square tests were used to determine associations between sociodemographic factors and outcomes. We used independent t-tests to compare mean ages of main outcome variables.

## Results

In total, 132 YEH participated in the study; among them, 10 reported HIV positive status and their data were not included in this analysis. Some of the youth chose not to complete the demographic portion of the survey and we removed those cases from analysis also. Among the 113 YEH in the analytic sample, the mean age was 21 years old, 52% identified as male, 36% female, 8% transgender, 5% other or non-binary, and nearly half (48%) identified as LGBTQIA (Table 1). Approximately two-thirds (69%) reported having health insurance and 66% reported experiencing food insecurity. Thirteen percent of the participants reported engaging in transactional sex. Injection drug use was rare (N=2), while 72% of participants reported condomless sex.

**Table 1.**
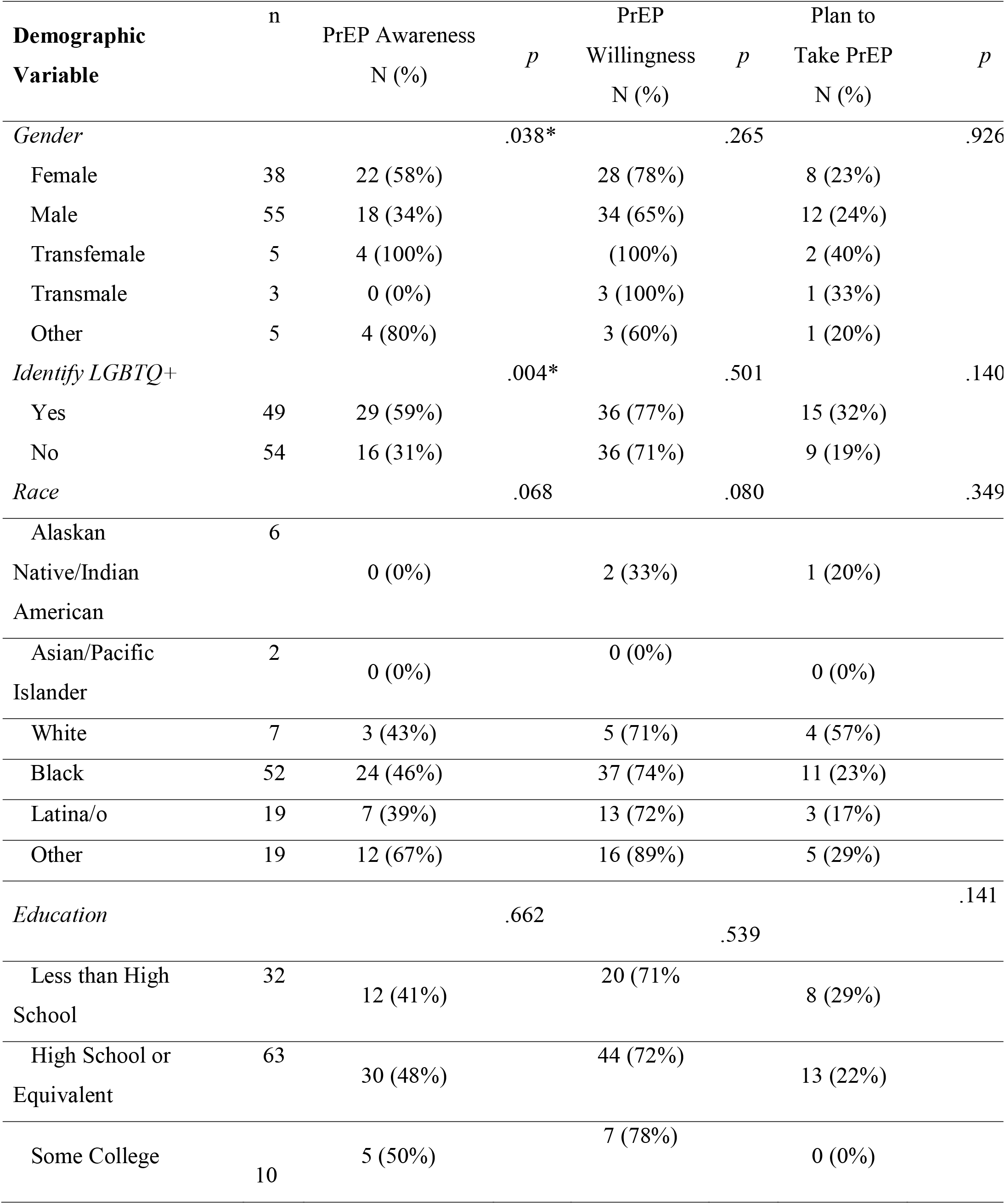
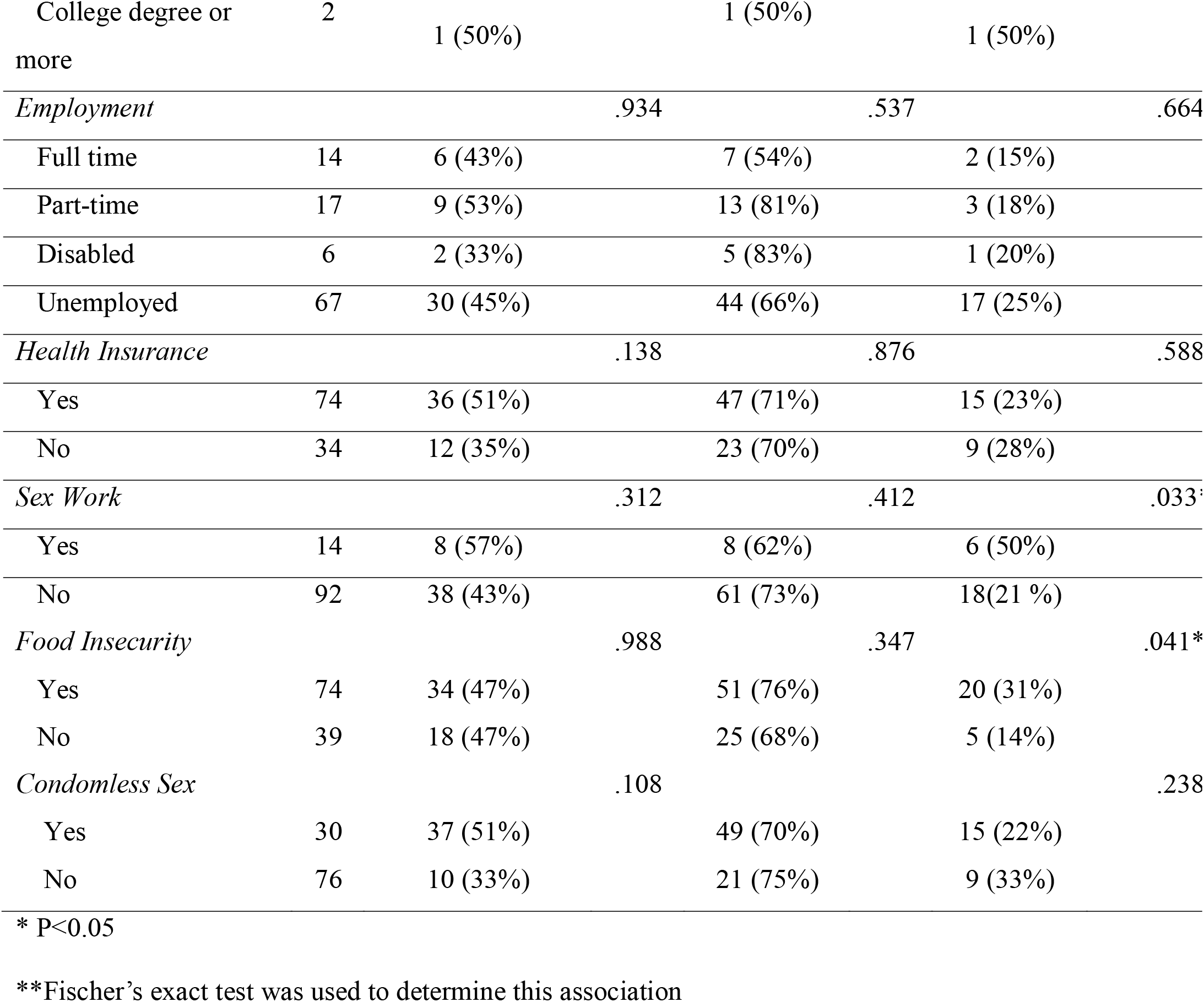
PrEP awareness, PrEP interest, and plans to take PrEP among youth experiencing homelessness in NYC (N=113).

Table 1 demonstrates bivariate associations between baseline characteristics and primary outcomes: PrEP awareness, willingness to take PrEP, and future plans to take PrEP. Overall, 47% reported awareness of PrEP before taking the survey. Female participants were more likely than male participants to be aware of PrEP (58% vs. 34%; *p =* .*038*). Participants who identified as LGBTQ+ were more likely to be aware of PrEP versus those who did not identify as LGBTQ+ (59% vs. 31%; *p =* .*004*). While 73% of the participants reported willingness to take PrEP, only 25% had a plan to take PrEP. Participants who reported food insecurity were more likely to have a plan to take PrEP versus those who did not report food inseucurity (31% vs. 14%; *p =* .*041*). Fischer’s exact test was used to determine the association between plans to take PrEP and food insecurity because there were cells containing less than 5 cases. Finally, YEH who reported transactional sex were more likely to have a plan to take PrEP than those who did not report transactional sex (50% versus 21%; *p =* .*033*). We conducted independent sample t-tests with age and each of our primary outcome variables. We did not find any significant differences based on age. The average age was 21 years old for PrEP awareness (t = -.213, *p =* .*416*), willingness to take PrEP (t = .723, *p =* .*236*), and plans to take PrEP (t = -.046, *p =* .*482*).

Only 11 participants (10%) had previously received a PrEP prescription, of which 5 reported actually taking it. Additionally, three participants reported taking PrEP without a prescription. The most commonly identified barrier to using PrEP was concern about side effects (N=22, 21%). Remembering to take a pill every day (N = 10, 10%), access to a provider to prescribe PrEP (N = 8, 8%), and access to a pharmacy to pick up PrEP (N = 8, 8%) were all rarely identified as barriers.

## Discussion and Conclusions

Almost no prior studies have addressed PrEP engagement among YEH, despite their exceedingly high risk of HIV. We examined PrEP engagement among YEH in NYC, finding low levels of PrEP awareness and future plans to take PrEP, despite high willingness to take it. Notably, our study included a more representative sample than the few prior investigations, which recruited few participants identifying as female, transgender or nonbinary, and were predominantly single-site studies. We surveyed YEH at 9 different sites, reaching a sample that included 31% female participants and 13% who identified as transgender or non-binary, more closely mirroring NYC’s YEH population.

Less than half of participants were aware of PrEP; notably, awareness was more likely among those identifying as LGBTQ+. Similarly, Storholm and colleagues found that LGBTQ+ identification was positively associated with PrEP engagement.^11^ This may be an indicator of successful targeted PrEP messaging to this key population for HIV prevention. Marketing PrEP to non-LGBTQ+ youth that are at risk is also essential, seeing that HIV risk is also exacerbated by housing instability or homelessness.^2^ There were no other significant differences significantly associated with PrEP awareness. Lack of knowledge of PrEP among YEH is likely a prominent barrier to PrEP engagement, and may reflect structural barriers associated with homelessness (e.g., lower levels of education and health literacy, poor access to medical care). To address lack of awareness, we suggest disseminating information about PrEP through YEH drop in centers and other networks, like social media. Flyers about PrEP might be shared or placed in common “hangout” locations in NYC.

It will also be important to improve access to PrEP in places where YEH are easily able to engage, such as drop in centers and primary care facilities. Community based organizations have successfully sent navigators into low resourced communities to introduce PrEP and other HIV prevention strategies like regular testing through mobile vans. Reaching people in the community experiencing homelessness, particularly YEH, can pose additional challenges because of the transient nature of homelessness. Integrating PrEP providers into drop in centers or inviting collaborations with patient or peer navigators at local hospital systems might also be effective ways to improve awareness. Interventions that are targeted toward this population should focus on educating YEH on the benefits of PrEP and connecting them to providers.

We observed high interest in taking PrEP, with over 70% of participants reporting willingness to take it. No sociodemographic characteristics were associated with willingness to take PrEP, and furthermore, there were few widely reported direct barriers to taking it. This may be because more than half of the sample did not know about PrEP. Concern about side effects was the most frequently reported barrier to regularly taking PrEP in our sample, reported by 20% of our participants. We should consider concerns about side effects and ensure that providers discuss this upfront and provide suggestions to manage side effects and the adjustment period. However, a study done by Mullins and Lehmann^12^ found that adolescents (albeit non-homeless) typically did not discontinue PrEP because of side effects. More research is needed to learn more about barriers to PrEP use among adolescents, particularly those who are experiencing homelessness.

Less than 30% of participants had future plans to take PrEP, despite high levels of interest. Future plans to take PrEP were more likely among respondents who reported food insecurity and who engaged in transactional sex. Food and housing insecurity are associated with higher risk for HIV infection in the literature.^2,7^ So, it is possible that participants who reported food insecurity were more aware of their risk, making them more likely to have plans to take PrEP. The sample had relatively few people with histories of transactional sex so this will require further investigation. There were no other significant demographic differences in plans to take PrEP, including having health insurance.

## Limitations

While we collected data from nine different sites frequented by YEH in NYC, and included a sample that is reflective of national YEH demographics, the study was conducted in one city in the United States, and results may not be generalizable to other geographic locations. Participants self-reported responses, which may have led to under-reporting of transactional sex and/or drug use as well as social desirability bias with respect to data on PrEP outcomes. Participation was anonymous, though, in order to minimize any self-report bias. Our data was cross-sectional, which prevents us from investigating causality. Finally, we collected data on LGBTQ+ individuals as one group, which overlapped with gender identity so targeted efforts might be needed to disaggregate gender and sexual orientation in future studies.

## Conclusions

We conducted an innovative study with implications for future research. We found that many YEH are not aware of PrEP, yet report high interest in using it. Future research should focus on how to improve awareness of and uptake of PrEP, potentially through community-based interventions that include tailored education for YEH about sexual health and PrEP. Additionally, longitudinal studies will be important to investigate causal mechanisms involved in PrEP engagement. Given the increasing burden of HIV among youth and YEH in particular, understanding how to reach this group and improve awareness and access to PrEP is critical to public health goals to end the HIV epidemic.

## Data Availability

All data produced in the present study are available upon reasonable request to the authors

## Declarations

The authors have no financial or non-financial interests to disclose.

## Notes

### Competing Interest Statement

The authors have declared no competing interest.

### Funding Statement

This study was funded by the PSC CUNY Foundation.

### Author Declarations

Approval was granted by the City University of New York Institutional Review Board. The CUNY Institutional Review Board granted a waiver of signed consent for this study, such that completion of the anonymous survey was considered implied consent to participate.

